# Size of societal volunteering predicts COVID-19 mortality

**DOI:** 10.1101/2022.03.31.22273238

**Authors:** Fritz Schiltz, Hans Van Remoortel, Hans Scheers, Philippe Vandekerckhove

## Abstract

**Introduction:** Different countries responded differently to the COVID-19 pandemic in terms of timing and stringency of measures, and in types of policies adopted. Typically, policy makers tried to balance the capacity of healthcare systems to take care of the ill (as determined by ICU capacity, availability of nurses, etc.), with safeguarding economic output (preventing total lockdown of the labor force, etc.). Later on, also a broader array of considerations such as impact on schooling or the need for social contact were taken into account to varying degrees.

The broad and relatively fast availability of data on healthcare and economic capacity, together with the political estimate that these were the most critical determinants for maintaining societal structure and compliance with the measures taken, in many countries prioritized decision-making. What received far less attention, in part due to the difficulty of obtaining reliable data in a timely manner, was the opposite question: to what extent do societal structures – besides healthcare and economic systems - contribute to a country’s resilience during catastrophes such as the pandemic? While it is commonly understood that the impact of a pandemic goes beyond its death count, perhaps the death count itself is impacted by the way societies are structured.

One example of such societal structure is the contribution of volunteers during the COVID-19 response. Volunteers may contribute to well-functioning societies in different ways, both through practical actions (e.g. knitting face masks) as by strengthening societal cohesion (e.g. encouraging fellow citizens to comply with measures). This paper quantifies the association between COVID-19 mortality and the size of societal volunteering, using the unique context of the COVID-19 crisis with its intensity, sudden onset and global spread.

## Methodology

### Study population

A ‘between countries’-comparison using international data, and an ‘in country’-comparison of the US using data on all 50 states was done. The international analysis was restricted to ‘high income’ countries^1^ and excluded smaller countries (less than 3 million inhabitants) to avoid including tax havens. Within this subset, 32 countries^2^ were retained based on availability of data on volunteering.

### Data collection

The dependent variable of interest is COVID-19 mortality, measured as the number of deaths due to COVID-19 per million inhabitants, from January 2020 until January 2022. The explanatory variable of interest is the size of societal volunteering. Different measures were sought to enable consistent analysis between the international and US states comparison. However, only very limited data is available on volunteering, and is typically context-dependent, with differing definitions, complicating comparisons across regions. Table A1 lists the 5 options finally considered. The choice was made to define volunteering as ‘organization-based’ volunteering, as per the definition of the International Labor Organization (ILO). The motivation for this choice is twofold. First, to the best of our knowledge, this definition is the only standardized measure that can be readily embedded in national labor force surveys.^3^ Second, this measure is – relative to other indicators - broadly and consistently adopted, making data comparable across regions. For example, the UN uses this definition as part of their *Satellite Account on Non-profit and Related Institutions and Volunteer Work*, and their *State of the World’s Volunteerism Report*. Eurostat is using a very similar measure for their reporting on *Social participation and integration statistics*, and the US Census Bureau surveys citizens on their volunteering activities within an organization or association in their *Current Population Survey Volunteers Supplement*.

For both international and US states comparison, data were collected for additional explanatory variables of COVID-19 mortality. Variables with available data for all selected countries and states include ‘general’ variables (GDP per capita, population density), variables related to the health care policy (health expenditure per capita, number of hospital beds per capita), COVID-19 policy interventions (stringency of COVID-19 measures implemented, vaccination rate), and COVID-19-specific predictors of mortality (Dessie, 2021) (prevalence of obesity, prevalence of smoking, share of population over 65 years old). In contrast to variables on volunteering, abundant data on these societal characteristics was publicly available for both international and US states analysis. Depending on the variable, sources of data can differ between the international analysis and the US analysis. An overview of the data sources per variable is presented in Table A2.

### Statistical analysis

Volunteering size as a predictor of COVID-19 mortality was analyzed in two steps for both the international and US states comparison. First, using a simple linear regression. Second, using a multiple linear regression model. Each explanatory variable was used in a simple regression to explain the variation in COVID-19 mortality. In case this relationship was significant (p-value<0.05), the variable was inserted in the multiple regression model.^4^ In addition, each control variable considered was compared to size of societal volunteering in its ability to predict COVID-19 mortality (r-squared, R^2^). All statistical analyses were performed using the software R (4.1.2).

Different assumptions were tested for both the simple and multiple models: (1) linearity; (2) normality of residuals; (3) multicollinearity (for the multiple regression only); (4) homoscedasticity; and (5) uncorrelatedness of independent variables with the error term. All 5 assumptions were confirmed for both models, with the exception of (4) in the simple model for the international analysis. In the results section, this model was hence estimated using a weighted linear regression to account for heteroscedasticity observed in the errors.

## Results

Figure 1 displays the association between size of societal volunteering and COVID-19 mortality, for the international and US states comparison, respectively. For both datasets, a significantly negative correlation exists (p-value<0.001), with size of societal volunteering explaining around 35% of observed variation in COVID-19 mortality (R^2^). Figure 2 compares the explanatory power of different variables. This figure shows that – apart from prevalence of smokers only in the international but not US states comparison – size of societal volunteering explains most of the observed variation in COVID-19 mortality across countries and between US states.

**Figure 1:**
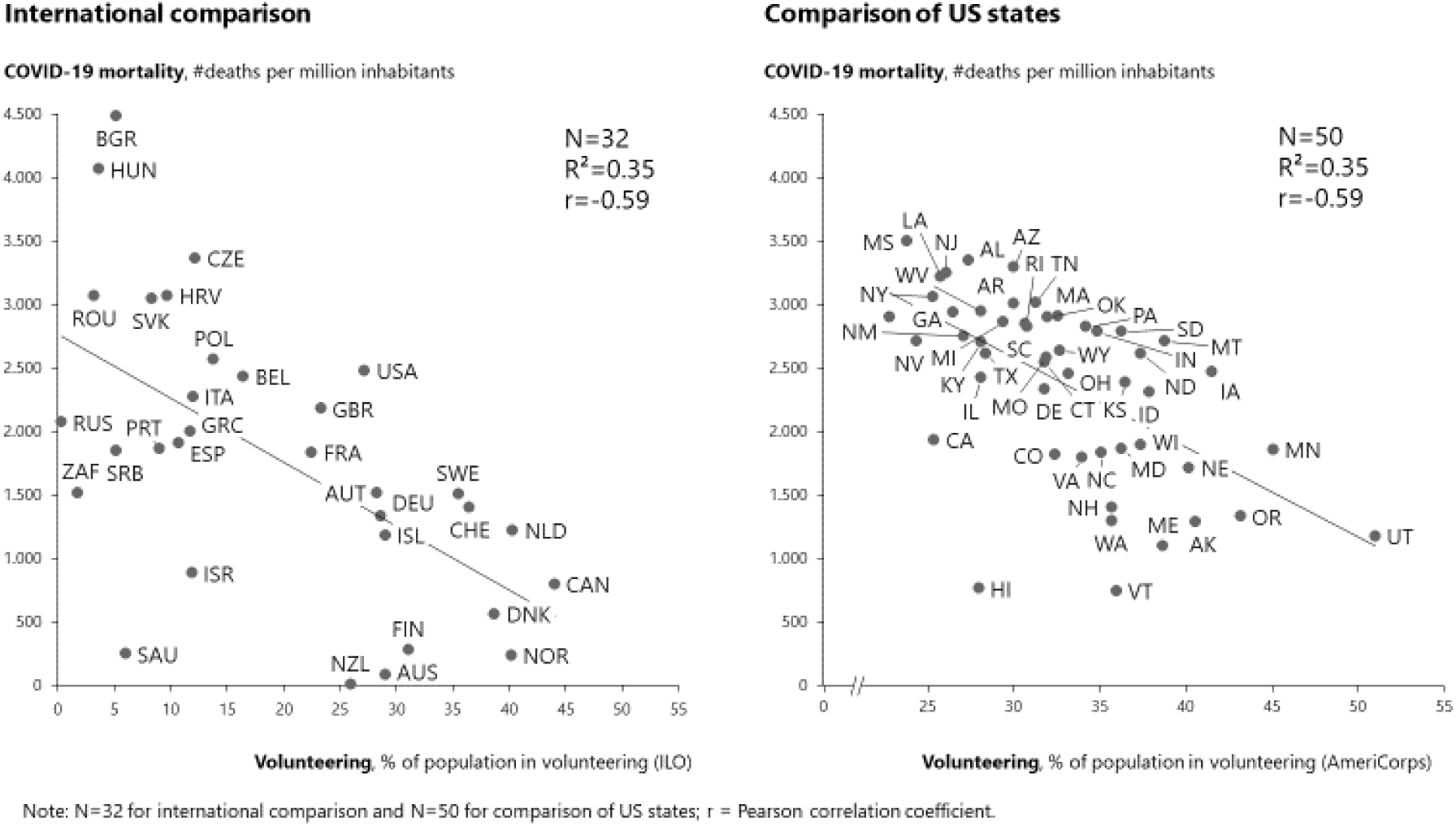
Societal volunteering and COVID-19 mortality.

**Figure 2:**
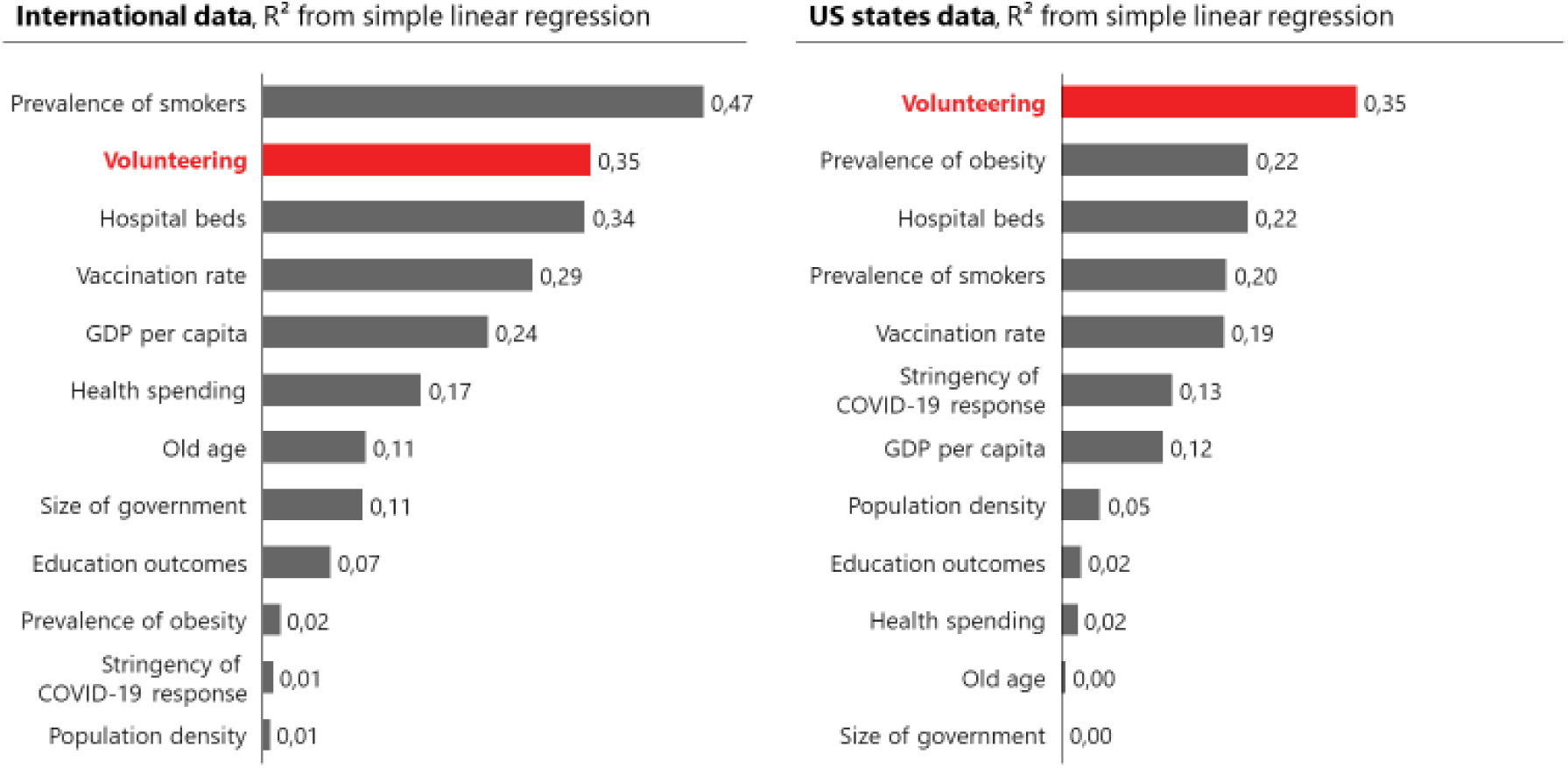
% of variance in COVID-19 mortality explained by different variables.

Table 1 depicts the results for the simple regression model (without control variables) and for the multiple regression model (including significant control variables as described). The coefficient of volunteering size is consistent in size between the simple and multiple regression analysis. In the multiple regression model, the coefficient equals -34,49 (international) and -75.37 (US states). Hence, for each percentage point increase in societal volunteering (% of population), the number of COVID-19 deaths per million inhabitants decreases with 34.49 and with 75.37, for the international and US states comparison, respectively. Due to the lower number of observations in the international comparison (N=32), the coefficient in the multiple regression model is not significant (p-value=0.09) whereas the multiple regression model for US states (N=50) remains significant (p-value<0.001). This difference in significance might be due to the lower number of observations in the international comparison (32 vs 50) or due to a stronger within-country effect (US states) compared to the cross-country effect (international comparison). However, under the constraints of the available data, both effects cannot be disentangled. For both study populations, adding control variables (such as % smokers, % fully vaccinated, and % obese) extends the explanatory power of the model (R^2^) to 0.53 (international comparison) and 0.59 (US states). In other words, more than half of the variance in COVID-19 mortality can be explained by the variance of the explanatory variables included in the model.

**Table 1:** Results of regression analysis with COVID-19 mortality as outcome variable.

|  | International comparison |  | US states comparison |  |
| --- | --- | --- | --- | --- |
|  | Simple | Multiple | Simple | Multiple |
| **Coefficient on size of societal volunteering** | -50.11 | -34.49 | -70.62 | -75.37 |
| **Standard error** | 12.65 | 19.67 | 13.90 | 11.83 |
| **p-value** | <0.001 | 0.09 | <0.001 | <0.001 |
| **R-squared (adjusted)** | 0.35 | 0.53 | 0.35 | 0.59 |
| **N** | 32 | 32 | 50 | 50 |
| Control variables included in multiple regression model for:<br>- international comparison: vaccination rate (%), hospital beds per 1,000 inhabitants (#), spending on health care per capita (USD), prevalence of smokers (%), GDP per capita (USD);<br>- US states comparison: vaccination rate (%), hospital beds per 1,000 inhabitants (#), obesity prevalence (%), prevalence of smokers (%), stringency index, GDP per capita (USD). |  |  |  |  |

As additional robustness check, we validated the multiple regression model by retaining the most significant variables in order to ensure a minimum number of observations per explanatory variable, thus restricting the number of explanatory variables to 1 for every 10 observations (i.e. countries or US states) (Harrell et al., 1984). In other words, we retained the 3 most significant variables for the international analysis (volunteering, hospital beds, and smokers) and the 5 most significant variables for the US states analysis (volunteering, vaccination rate, hospital beds, obesity, and stringency of the policy response). Regression results from this robustness check indicate a smaller coefficient for volunteering in the international analysis (−22.7) with equivalent significance as in the multiple model (p-value=0.08), while the regression results for US states are equivalent to the multiple regression model both in size and significance of the coefficient on volunteering (−75.4, p-value<0.001).

## Discussion

Our results show that societies with more volunteering are less severely affected by COVID-19. Although no causal conclusions can be drawn from these observational data, this effect is both statistically significant and clinically meaningful. Assuming the associations are causal and all countries and states perform at ‘best in class’-level of societal volunteering (Canada and Utah, respectively), the theoretical number of deaths avoided amounts to 41% (or 960,000) deaths for the international countries and 63% (or 510,000) for the US. The associations remain intact even after adding two layers of robustness checks (different regression models and different datasets). It is noteworthy that, apart from prevalence of smokers (and this only in the international comparison), no variable better predicts COVID-19 mortality than size of societal volunteering. This includes key predictors such as ‘high risk groups’ (e.g. people above 65 years old, obesity), expenditure on health care, and population density. It should be noted that when re-calculating COVID-19 mortality starting from the launch of the global vaccination campaign (December 2020) rather than the start of the pandemic, vaccination rate outperforms size of societal volunteering (r=0.39 vs r=0.35) but this only in the international comparison. However, even when including vaccination rate as a control variable, volunteering remains a significant predictor (p-value=0.02) of COVID-19 mortality in this period (December 2020-January 2022).

Due in part to the COVID-19 crisis with its disruptions of supply chains and predatory price setting for critical goods, the role of the state, after decades of decline, is emphasized again. Whether in innovation (e.g. vaccine development) or in strategic industries (e.g. face masks, ventilators, etc.), the public sector is once again strengthened vis-a-vis the private sector. Mintzberg (2015) theorized that a society cannot successfully exist if not based on three balanced pillars: a respectable and respected government (that sets and enforces the rules), a dynamic private sector (that innovates to pursue commercial success within the rules set), and a third sector based on citizens who voluntarily band around wrongs or oversights not (yet) addressed by the public or private sector. In two different datasets on third sector size (Salamon, Sokolowski & Haddock, 2017) (Salamon & Sokolowski, 2018), a strong correlation between size of societal volunteering and of third sector is present (r=0.69^5^ and r=0.64^6^ respectively). Hence, our findings suggest that the duality ‘private versus public sector’, often at the heart of the political debate, might oversimplify reality by ignoring third sector impact, thus confounding both analysis and potential effective solutions.

The results presented here are consistent with recent evidence that interpersonal trust and trust in government are key predictors for cross-national variation in the number of infections per capita (COVID-19 National Preparedness Collaborators, 2022). Although the description of causal mechanisms is outside the scope of this study, a preliminary analysis indicates that countries or states with more volunteering are also places where interpersonal trust is highest (r=0.74 for countries; r=0.60 for US states), using data from the World Values Survey (international), and the General Social Survey (US states). Whether cause or effect, more volunteering and trust correlate with better pandemic preparedness, with volunteering being an actionable target for policy makers.

The lack of comparable data limits studies such as ours to countries and states where standardized data is available on (a proxy of) third sector size. When collecting data for this study, it was striking to observe the abundance of data available for the private and public sector (e.g. Ease of Doing Business Index – World Bank, Digital Government Index - OECD), whereas almost no standardized data are collected on the third sector. The increased accuracy of the regression model for US states versus international comparison, because of the higher number of observations (N=50 vs N=32), highlights the research potential when more standardized data is available. Further research requires systematic data collection and (quasi-)experimental study designs. In order to strengthen the empirical basis and expand to other proxy’s for third sector, it is necessary to 1) systematically use ILO-definitions to measure the size of societal volunteering; 2) define international standards to register other third sector data; 3) systematically collect data on both.

## Data Availability

All data produced in the present study are available upon reasonable request to the authors

## Supplemental Appendix

**Table A1:** Comparison of different available indicators for size of societal volunteering.

| Indicator | Source | Available for US states analysis | Available for international analysis |
| --- | --- | --- | --- |
| % of population (16+) in formal voluntary activities | Eurostat | NO | YES (EU only) |
| Civil society workforce (volunteers) as % of economically active population | Salamon, Sokolowski, & Haddock (2017) | NO | YES |
| Direct volunteers as % of national employment | Salamon & Sokolowski (2018) | NO | YES (EU only) |
| % of population (16+) in volunteering, organization-based | International Labor Organization / AmeriCorps | YES | YES |
| % of population (16+) in volunteering, total | International Labor Organization | NO | YES |

**Table A2:** Data sources.

| Indicator | International comparison |  | US states comparison |  |
| --- | --- | --- | --- | --- |
|  | Indicator | Source | Indicator | Source |
| COVID-19 mortality | # deaths per million inhabitants | Johns Hopkins University | # deaths per million inhabitants | Johns Hopkins University |
| Size of societal volunteering | % of population (16+) in volunteering, organization-based | International Labor Organization | % of residents in volunteering (~ILO definition) | Corporation for National and Community Service |
| GDP per capita | Per capita income (USD) | World Bank | Per capita income (USD) | US Census Bureau |
| Population density | Population per square kilometer | World Bank | Population per square mile | US Census Bureau |
| Size of government | Government expenditure as % of GDP | World Bank | State and local spending as % of GSP | US Census Bureau |
| Education outcomes | PISA score (average math, reading, science) | OECD | NAEP score (math 8th grade) | National Center for Education Statistics |
| Vaccination rate | % of people fully vaccinated | Johns Hopkins University | % of people fully vaccinated | US Centers for Disease Control and Prevention |
| Health spending | Health expenditure per capita (USD) | World Bank | Health expenditure per capita (USD) | US Centers for Medicare & Medicaid Services |
| Hospital beds | Hospital beds per 1,000 population | World Bank | Hospital beds per 1,000 population | American Hospital Association |
| Stringency of COVID-19 response | Average of stringency index | University of Oxford | Average of stringency index | University of Oxford |
| Prevalence of obesity | Obesity rate (% of people with BMI $\geq 30$) | World Health Organization | Obesity rate (% of people with BMI $\geq 30$) | US Centers for Disease Control and Prevention |
| Prevalence of smokers | Tobacco use (% of 18+) | World Health Organization | Current smokers (% in 18+ population) | US Centers for Disease Control and Prevention |
| Old age | % of population 65 and above | World Bank | % of population 65 and above | US Census Bureau |
| Note: For each variable, data for the latest available year was collected. In case data was available for 2020 or 2021, data was collected for 2019, to avoid interference with COVID-19 mortality. All data is publicly available, and the master dataset is available upon request from the authors. |  |  |  |  |

**Table A3:**
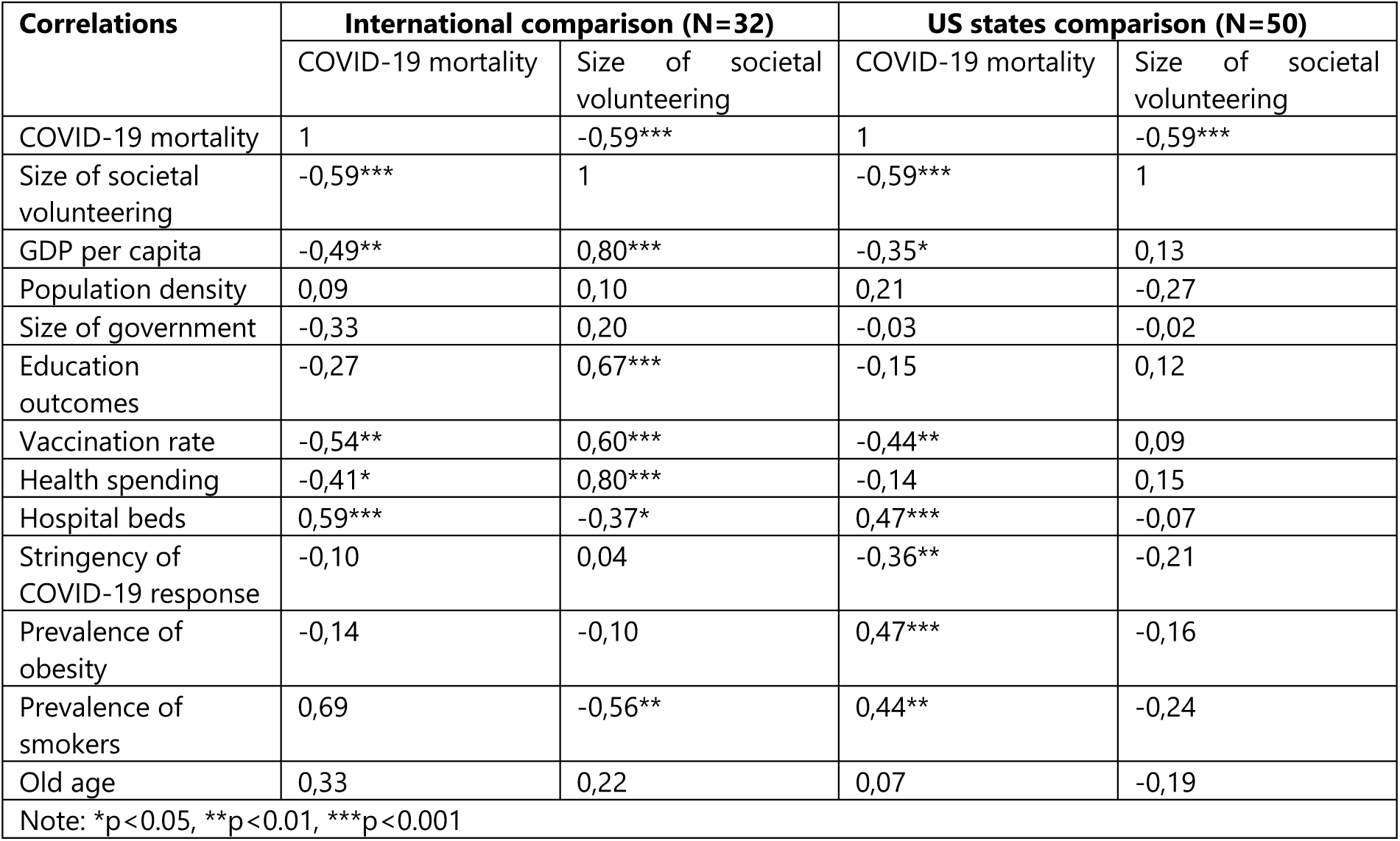
Pearson correlation between outcome variable (COVID-19 mortality), explanatory variable of interest (size of societal volunteering), and other explanatory variables.

| Correlations | International comparison (N=32) |  | US states comparison (N=50) |  |
| --- | --- | --- | --- | --- |
|  | COVID-19 mortality | Size of societal volunteering | COVID-19 mortality | Size of societal volunteering |
| COVID-19 mortality | 1 | -0,59*** | 1 | -0,59*** |
| Size of societal volunteering | -0,59*** | 1 | -0,59*** | 1 |
| GDP per capita | -0,49** | 0,80*** | -0,35* | 0,13 |
| Population density | 0,09 | 0,10 | 0,21 | -0,27 |
| Size of government | -0,33 | 0,20 | -0,03 | -0,02 |
| Education outcomes | -0,27 | 0,67*** | -0,15 | 0,12 |
| Vaccination rate | -0,54** | 0,60*** | -0,44** | 0,09 |
| Health spending | -0,41* | 0,80*** | -0,14 | 0,15 |
| Hospital beds | 0,59*** | -0,37* | 0,47*** | -0,07 |
| Stringency of COVID-19 response | -0,10 | 0,04 | -0,36** | -0,21 |
| Prevalence of obesity | -0,14 | -0,10 | 0,47*** | -0,16 |
| Prevalence of smokers | 0,69 | -0,56** | 0,44** | -0,24 |
| Old age | 0,33 | 0,22 | 0,07 | -0,19 |
| Note: *p<0.05, **p<0.01, ***p<0.001 |  |  |  |  |

## Footnotes

1 At the time of writing, this cutoff stood at 12,696USD GNI per capita, see https://datahelpdesk.worldbank.org/knowledgebase/articles/906519#High_income

2 Australia, Austria, Belgium, Bulgaria, Canada, Croatia, Czechia, Denmark, Finland, France, Germany, Greece, Hungary, Ireland, Israel, Italy, Netherlands, New Zealand, Norway, Poland, Portugal, Romania, Russia, Saudi Arabia, Serbia, Slovakia, South Africa, Spain, Sweden, Switzerland, United Kingdom, United States.

3 For details on the data collection and definition of this indicators, see ILO’s manual for the add-on module for labor force surveys: https://www.ilo.org/wcmsp5/groups/public/---dgreports/---stat/documents/publication/wcms_789950.pdf

4 Table A3 lists the Pearson correlations between the outcome variable (COVID-19 mortality), the explanatory variable of interest (size of societal volunteering), and other explanatory variables.

5 Compared to international analysis, Bulgaria, Croatia, Greece, Saudi Arabia, and Serbia are not included here.

6 When restricting the correlation to the subset of 22 European countries with available data.

